# Causal contribution of malignancy to anomalous baroreflex functionality in patients with cancer: An overlooked aspect of cardio-oncology

**DOI:** 10.1101/2020.10.28.20221044

**Authors:** Chi Fang, Ching-Yi Tsai, Chih-Yen Chien, Samuel H.H. Chan

## Abstract

The current trend in cardio-oncology places major emphasis on circulatory toxicity induced by cancer therapy. Whether malignancy itself is a direct contributing factor to cardiovascular dysfunctions and baroreflex dysregulation in patients with cancer, however, has rarely appeared in literature. The present study addressed this largely overlooked aspect of cardio-oncology by evaluating blood pressure, heart rate and baroreflex functionality before and after curative surgery in patients with oral cavity squamous cell carcinoma (OSCC). We found that these patients exhibited reduced baroreflex-mediated sympathetic vasomotor tone and augmented cardiac vagal baroreflex, and such inherent anomalies were readily reversed to levels of healthy controls after surgical removal of the primary tumor. It is concluded that by being more prone to hypotension and bradycardia, anomalous baroreflex functionality causally induced by malignancy predisposes patients with cancer to detrimental cardiovascular abnormalities that may be further exacerbated by cancer therapy.

## Introduction

Historically, there have been continuous interests in the relationship between cardiovascular diseases and malignancy. With the establishment of the disciplinary of cardio-oncology, the current emphasis is primarily with circulatory toxicity induced by cancer therapy (Lenneman and Sawyer, 2016; Minasian et al., 2019). Whether malignancy itself is a direct contributing factor to cardiovascular dysfunctions and baroreflex dysregulation in patients with cancer, however, has rarely appeared in literature. The present study addressed this largely overlooked aspect of cardio-oncology by evaluating blood pressure (BP), heart rate (HR) and baroreflex functionality before and after curative surgery in patients with oral cavity squamous cell carcinoma (OSCC). Our rationales are fourfold. BP and HR are the two most common cardiovascular measurements in clinical practice; baroreflex is the most fundamental mechanism in cardiovascular autonomic regulation; global incidence of OSCC is on the rise (Thomson, 2018); and tumor excision provides the most direct assessment of causation.

## Methods

Our study population comprised patients with confirmed OSCC who were scheduled between January and October, 2016 for curative surgical removal of the primary tumor. They were classified according to the American Joint Committee on Cancer (AJCC) staging manual (Amin et al., 2017). Patients who have been receiving antihypertensive medication were excluded to eliminate this confounding factor. Based on this exclusion criterion, results from 36 patients (mean age = 55.4 years) formed the basis of this report. Normal healthy controls (n = 19; mean age = 34.9 years) were research staff from the Institute for Translational Research in Biomedicine.

Our study protocol followed the ethical guidelines of the Declaration of Helsinki, and Institutional Review Board approval (104-6534B) was obtained from Chang Gung Medical Foundation, Taiwan. All participants gave written informed consent. One day before and 7-10 days after surgery, BP was recorded noninvasively from the index finger by photoplethysmography (CNAP monitor 500, CNSystems Medizintechnik GmbH, Reininghausstraße, Austria). On-line digitized BP signals (Notocord, Croissy-Sur-Seine, France) were first analyzed by an arterial blood pressure analyzer (APR31a, Notocord) to obtain systolic BP (SBP), mean BP (MAP), HR, and pulse interval (PI). SBP signals were subsequently subjected to spectral analysis (SPA10a, Notocord) to detect the power density of the low-frequency (0.04-0.15 Hz) component in the SBP spectrum, an index for spontaneous baroreflex-mediated sympathetic vasomotor tone (Li et al., 2001; Tsai et al., 2019). To evaluate the cardiac vagal baroreflex, we used a baroreflex sequence analyzer (BRS10a, Notocord) to determined baroreflex sensitivity (BRS) based on detection of spontaneous sequences of consecutive increases or decreases in SBP and the associated changes in HR (Tsai et al., 2019).

## Results

The key finding of this study (**Figure 1**) is that, as a group, patients with OSCC already exhibited significantly reduced baroreflex-mediated sympathetic vasomotor tone under a condition when BP was comparable to healthy controls. At the same time, significantly augmented cardiac vagal baroreflex was seen concomitantly with reduction in HR. Intriguingly, the alterations in both arms of baroreflex and HR were significantly reversed to the level of healthy controls after surgical removal of OSCC. Breaking down the patients by stages of severity (**Figure 1**) revealed that whereas the above observations on baroreflex-mediated sympathetic vasomotor tone were only demonstrated in Stage 0-II patients, those on cardiac vagal baroreflex and HR were seen only in Stage III-IV patients.

**Figure 1.**
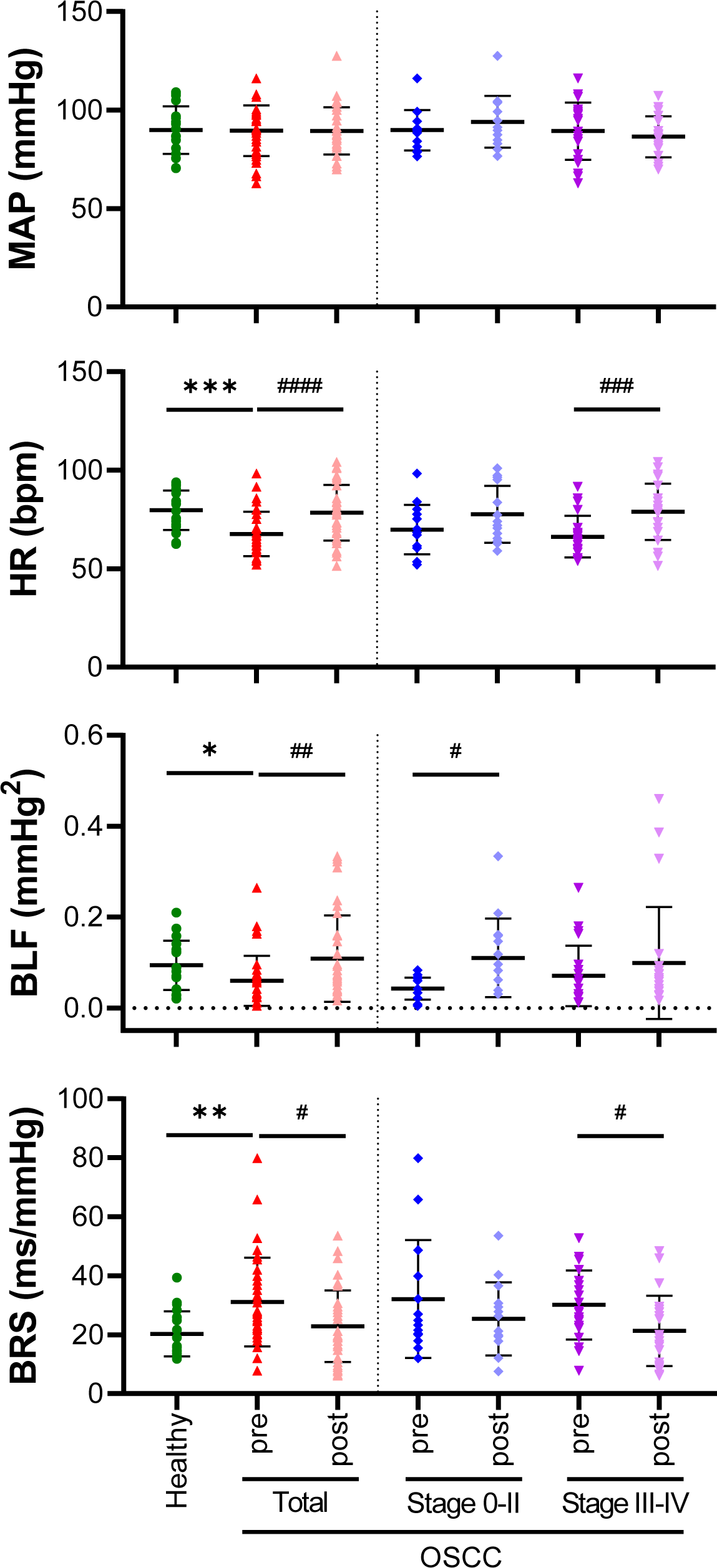
Mean arterial pressure (MAP), heart rate (HR) and both arms of baroreflex (BLF and BRS) in patients with squamous cell carcinoma (OSCC) before and after curative surgery. Values are mean ± SD of 36 patients with OSCC (14 patients under Stage 0-II, 22 patients under Stage III-IV) and 19 healthy subjects. With the exception of MAP (P = 0.7716), one-way ANOVA (F_6,156_) revealed statistically significant differences between the 7 group means for HR (P < 0.0001), BLF (P = 0.0382) and BRS (P = 0.0064). ^*^P < 0.05, ^**^P < 0.01 or ^***^P < 0.001 versus healthy control group in Student’s *t*-test; and ^#^P < 0.05, ^##^P < 0.01, ^###^P < 0.001 or ^####^P < 0.0001 versus corresponding pre-surgery group in paired Student’s *t*-test.

## Discussion

Our data revealed that the functionality of both the baroreflex-mediated sympathetic vasomotor and cardiac vagal baroreflex in patients with OSCC is anomalous even before manifestation of significant changes in BP. More importantly, we demonstrated that the primary tumor is causally related to this inherent baroreflex dysregulation.

Based on results from topographic analysis of the baroreflex neural circuits using diffusion tensor imaging, we proposed recently (Tsai et al., 2019) that the basal baroreflex-mediated sympathetic vasomotor tone is sustained by tonic inhibitory output from the nucleus tractus solitarii (NTS) where the baroreceptor afferents terminate to the rostral ventrolateral medulla (RVLM), which in turn lessens the tonic excitatory action of these premotor sympathetic neurons on vasomotor tone. Likewise, the basal cardiac vagal baroreflex activity is sustained by a tonic excitatory output from the NTS to the nucleus ambiguus (NA), which in turn exerts a tonic inhibitory action on the heart via the vagus nerve. Speculatively, an increase in the tonic inhibitory influence of the NTS on RVLM and an augmentation of the tonic excitatory influence of the NTS on NA in response to the same degree of baroreceptor activation in the healthy controls will result in the observed anomalous baroreflex functionality in patients with OSCC.

Controlling risk factors of cardiovascular diseases is stipulated to reduce the risk of cancer (Kiene et al., 2016). However, Coumbe and Groarke (2018) reviewed that the contribution of cardiovascular autonomic dysfunction to overall morbidity and mortality in cancer survivors has largely been overlooked. The presence of cancer in combination with reduced heart rate variability is also noted to be associated with shorter survival time (Guo et al., 2015). In terms of pathophysiology, patients with reduced baroreflex-mediated sympathetic vasomotor and augmented cardiac vagal baroreflex are more prone to hypotension and bradycardia. As such, it is conceivable that the inherent presence of anomalous baroreflex functionality would predispose patients with OSCC to detrimental cardiovascular abnormalities that may be further exacerbated by cancer therapy. Whether this causal relationship is applicable to all forms of cancer, and exactly how the primary tumor contributes to baroreflex dysfunction, particularly in its differential influence on baroreflex-mediated sympathetic vasomotor and cardiac vagal baroreflex that is dependent on the severity of malignancy, await further investigation. In this regard, using atherosclerosis as an illustrative example, intracellular signaling cascades that lead to inflammation have been proposed (Libby and Kobold, 2019) to be a pathophysiological commonality between cancer and cardiovascular diseases.

## Data Availability

Reasonable requests for data referred to in the manuscript will be honored by communication with the corresponding author.

## References

Amin M, Edge S, Greene F. AJCC Cancer Staging Manual. 8th edition. New York, NY: Springer, 2017.

Coumbe BGT, Groarke JD. Cardiovascular autonomic dysfunction in patients with cancer. Curr Cardiol Rep. 2018;20:69.

Guo Y, Koshy S, Hui D, et al. Prognostic value of heart rate variability in patients with cancer. J Clin Neurophysiol. 2015;32:516–520.

Kiene RJ, Prizment AE, Blaes A, Konety SH. Shared risk factors in cardiovascular disease and cancer. Circulation. 2016;133:1104–1114.

Lenneman CG, Sawyer DB. An update on cardiotoxicity of cancer-related treatment. Circ Res. 2016;118:1008–1020.

Li PL, Chao YM, Chan SHH, Chan JYH. Potentiation of baroreceptor reflex response by heat shock protein 70 in nucleus tractus solitarii confers cardiovascular protection during heatstroke. Circulation. 2001;103:2114–2119.

Libby P, Kobold S. Inflammation: a common contributor to cancer, aging, and cardiovascular diseases - Expanding the concept of cardio-oncology. Cardiovasc Res. 2019;115:824–829.

Minasian LM, Dimond E, Davis M, et al. The evolving design of NIH-funded cardio-oncology studies to address cancer treatment-related cardiovascular toxicity. J Am Coll Cardiol CardioOnc. 2019;1:105–113.

Tsai CY, Poon YY, Chan JYH, Chan SHH. Baroreflex functionality in the eye of diffusion tensor imaging. J Physiol. 2019;597:41–55.

Thomson, PJ. Perspectives on oral squamous cell carcinoma prevention - proliferation, position, progression and prediction. J. Oral Pathol Med. 2018;47:803–807.

